# Persisting Chemosensory Impairments in 366 Healthcare Workers Following COVID-19: An 11-Month Follow-up

**DOI:** 10.1101/2021.12.20.21268066

**Authors:** Nicholas Bussiere, Jie Mei, Cindy Levesque-Boissonneault, Mathieu Blais, Sara Carazo, Francois Gros-Louis, Robert Laforce, Gaston De Serres, Nicolas Dupre, Johannes Frasnelli

## Abstract

**Background and Objectives:** Olfactory and gustatory dysfunctions (OD, GD) are prevalent symptoms following COVID-19 and persist in 6%-44% of individuals in the first months after the infection. As only few reports have described their prognosis more than 6 months later, the main objective of this study was to assess the prevalence of OD and GD 11 months after COVID-19. We also aimed to determine test-retest reliability of subjective chemosensory ratings for the follow-up of chemosensory sensitivity, as this measure is often used for remote follow-up.

**Methods:** Inclusion criteria included a PCR-confirmed SARS-CoV-2 infection; exclusion criteria were the presence of other respiratory infections and chronic sinusitis. To assess whether OD and GD had changed compared to pre-pandemic levels, we designed an observational study and distributed an online questionnaire assessing quantitative chemosensory function to healthcare workers 5 and 11 months after COVID-19. Specifically, we assessed olfaction, gustation, and trigeminal sensitivity (10-point visual analog scale) and function (4-point Likert scale) separately. We further assessed clinically relevant OD using the Chemosensory Perception Test, a psychophysical test designed to provide a reliable remote olfactory evaluation. Qualitative chemosensory dysfunction was also assessed.

**Results:** We included a total of 366 participants (mean age of 44.8 years old (SD: 11.7)). They completed the last online questionnaire 10.6 months (SD: 0.7) after the onset of COVID-19 symptoms. Of all participants, 307 (83.9%) and 301 (82.2%) individuals retrospectively reported lower olfactory or gustatory sensitivity during the acute phase of COVID-19. Eleven months later, 184 (50.3%) and 163 (44.5%) indicated reduced chemosensory sensitivity, 32.2% reported impairment of olfactory function while 24.9% exhibited clinically relevant OD. Three variables predicted OD at follow-up, namely chest pain and GD during COVID-19 and presence of phantosmia at 5 months. Olfactory sensitivity ratings had a high test-retest reliability (intraclass correlation coefficient: 0.818 (95% CI: 0.760 - 0.860))

**Discussion:** This study suggests that chemosensory dysfunctions persist in a third of COVID-19 patients 11 months after COVID-19. Subjective measures have a high test-retest reliability and thus can be used to monitor post-COVID-19 OD. OD appears to be a common long-term symptom of COVID-19 important to consider when treating patients.

## Introduction

The COVID-19 pandemic has been ongoing for over two years, and many advances have been made to further understand its pathogenesis and treatment. The high rate of post-COVID-19 olfactory dysfunction (OD) has brought interest to the field of post-viral OD, yet many questions remain unanswered regarding the duration and pathophysiology of this symptom^1^. Even before the pandemic, viral infections of the upper respiratory tract (URTI) were known to be a major cause of OD^2^. For patients with persisting OD, impairment may be quantitative (e.g., anosmia, hyposmia) and/or qualitative (e.g., parosmia, phantosmia)^3^. Both forms of OD may significantly decrease quality of life (QoL), and this impact gets worse as OD persists^4^.

Initial presentation of COVID-19-induced OD is now well described: sudden loss affecting 50% to 75% of infected individuals^5^. For most individuals, quantitative OD is predominant, but some have reported parosmia (altered perception of real stimuli) and/or phantosmia (perception of odor in absence of stimuli) in the acute phase^6, 7^. Some individuals also present with other chemosensory alterations, such as gustatory dysfunction (GD; altered ability to taste sweet, sour, salty, bitter or umami) or trigeminal dysfunction (TD; altered ability to perceive spiciness, freshness, carbonation)^8-10^. Although most patients do recover within weeks from these dysfunctions, many remain symptomatic and their condition evolves into long-haul COVID-19^11, 12^. Six months after onset of symptoms, 5% to 60% of patients suffer from persistent OD and 10 to 35% have persistent GD^13-15^. Few studies reported prevalence of TD, although a recent study which used psychophysical tests found a correlation between OD and TD at 6 months following a confirmed SARS-CoV-2 infection^16^. This study could not establish a prevalence of trigeminal dysfunction due to their testing methods. Moreover, prevalence of parosmia increases with time in COVID-19 patients, which is in line with the theory that qualitative dysfunctions following viral infection could result from faulty regeneration of olfactory sensory neurons^17-19^. Although many patients with long-haul COVID-19 have persistent OD, very few studies describe the prognosis for such individuals past 6-months after symptom onset.

As described in a previous report, 52%, 42% and 23% of a cohort of healthcare workers with mild COVID-19 experienced OD, GD, and TD respectively at 5 months after onset of COVID-19^20^. For other viruses, recovery of postviral OD can be expected in up to 80% one year following onset of infection^21^. Therefore, long-term follow-up of patients is necessary to further understand the evolution of post-COVID-19 OD and eventually offer resources for clinicians and patients alike.

We followed up the same cohort of healthcare workers with a PCR confirmed SARS-CoV-2 infection between 12 February and 11 June 2020, who consented to fill an online questionnaire and to self-report measurements from the Chemosensory Perception Test (CPT), a novel and easy to use olfactory test. Data for the 5-month follow-up questionnaire from this cohort is published^20^. This present manuscript therefore compares data obtained at 5 and 11 months collected from the same cohort^20^.

We had three specific objectives for this study: first, we aimed to determine the percentage of individuals who experience OD, GD, or TD 11 months after COVID-19. To do so, we used three different approaches to grasp different aspects of chemosensory dysfunction. Specifically, we assessed (a) the proportion of participants who indicated chemosensory sensitivity below levels prior to the infection (persistent reduction of chemosensory sensitivity); (b) the proportion of participants who indicated chemosensory function was much or a bit worse than before the infection (impairment of chemosensory function); (c) the proportion of participants who had a result indicative of dysfunction in a semi-objective test (clinically relevant OD). Second, we aimed at identifying the factors that best predict olfactory dysfunction 11 months after COVID-19. Third, we aimed to determine the test-retest reliability of subjective chemosensory ratings.

## Methods

### Standard protocol approvals, registrations, and patient consents

This study was reviewed and approved by the research ethics board of the CHU de Québec – Université Laval (MP-20-2021-5228) and all protocols were reviewed by an independent Scientific Review Committee. All participants provided an online informed consent prior to participation. The study received funding from the Fonds de recherche du Québec-Santé (FRQS). No compensation or incentive was offered to participants.

### Participants

Healthcare workers with a positive SARS-CoV-2 PCR test were recruited among those who had completed the initial online questionnaire at 5 months after onset of symptoms. Inclusion criteria were (1) completed the follow-up online questionnaire (2) did not report other respiratory diseases (bacterial or viral infection) within 2 weeks prior to questionnaire completion, chronic sinusitis, or traumatic brain injury, and (3) did not have a symptomatic SARS-CoV-2 reinfection (Figure 1).

**Figure 1.**
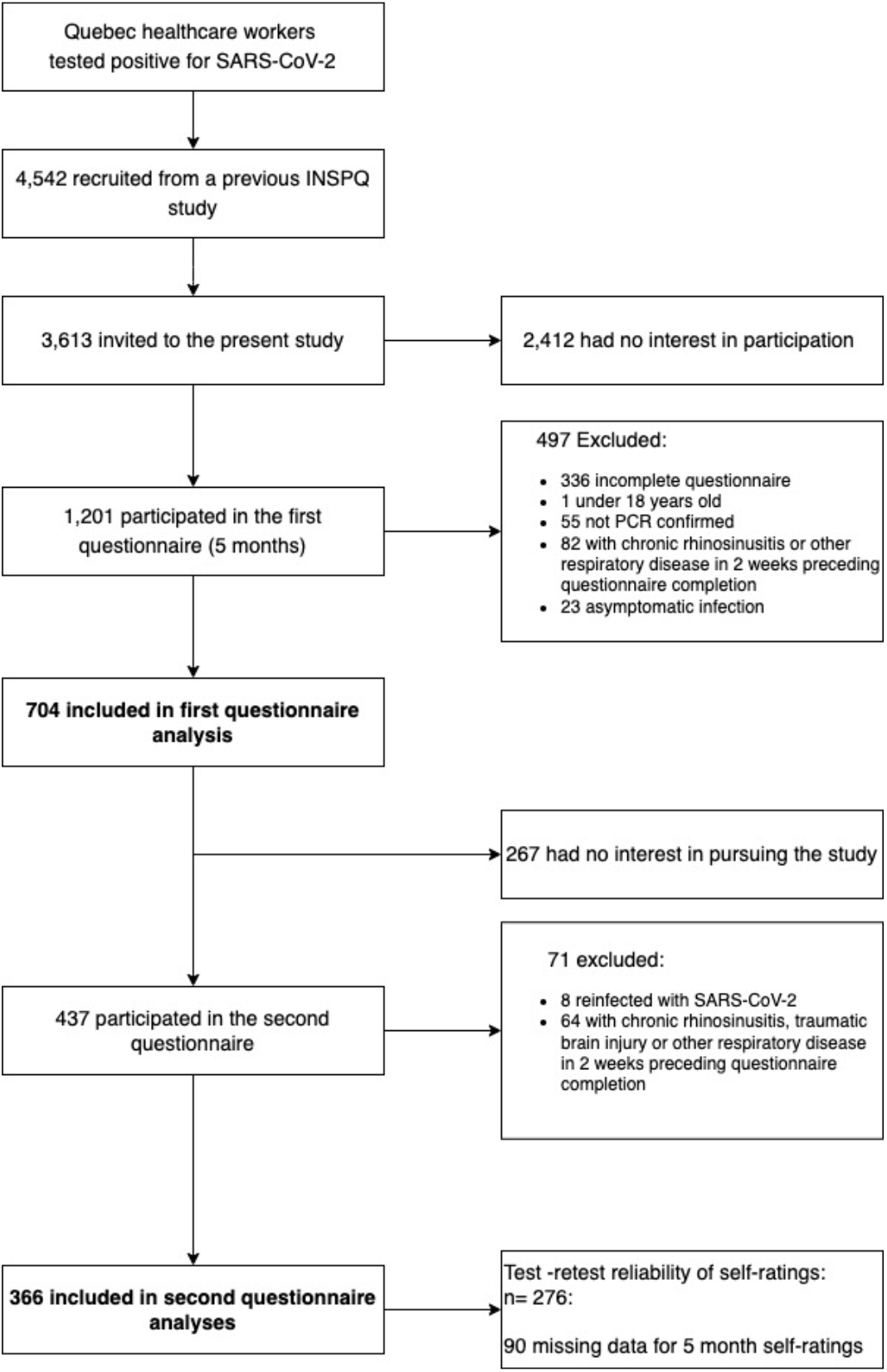
Participant inclusion flow chart. INSPQ : Institut national de santé publique du Québec

### Study design

Data were collected from March 1 to April 27, 2021. Up to four attempts were made via email to reach and recruit potential participants.

### Online questionnaire

All participants were invited to complete an online questionnaire as introduced earlier ^20^. The questionnaire comprised items on chemosensory self-assessment including demographic data, changes to medical history (based on the follow-up questionnaire of the Global Consortium on Chemosensory Research^8, 22^). Finally, we remotely administered the CPT, a semi-objective test for chemosensory function^20^ (eAppendix 1).

### Chemosensory self-assessment

Here, participants were asked to self-evaluate and report their olfactory *(i.e., the ability to perceive the smell of flowers, soap, or garbage but not the flavor of food in the mouth)*, gustatory (*i.e., the ability to perceive sweetness, sourness, saltiness, bitterness in the mouth*), and trigeminal (*i.e., the ability to perceive the spiciness of chili peppers, the cooling of menthol and the carbonation in soda*) sensitivity using a 10-point visual analog scale (VAS) at four time points: (T0) before SARS-CoV-2 infection (i.e. baseline), (T1) during SARS-CoV-2 infection, (T2) at initial questionnaire completion (approximately 5 months after SARS-CoV-2 infection) and (T3) at last questionnaire completion (approximately 11 months after SARS-CoV-2 infection). This allowed us to determine if the individual subjectively exhibited full recovery; when a participant rated their current (at T3) ability to be lower than their self-evaluation at baseline (at T0; i.e.: T3 < T0), they were classified as exhibiting (a) persistent reduction of chemosensory sensitivity. In addition, these data also allowed us to compare scores from one timepoint to another (i.e.: T0 vs T2 vs T3).

Participants were then asked to compare their current (at T3) chemosensory function with those prior to the SARS-CoV-2 infection (T0), for which they may choose one of the following statements: (1) *much worse*, (2) *a bit worse*, (3) *the same*, (4) *a bit better*, and (5) *much better*. Participants from categories (1) and (2) were classified as exhibiting (b) impairment of chemosensory function. For participants with self-reported OD (GD), we also collected information about the presence of current parosmia/phantosmia (alterations in the 5 taste qualities, i.e., sweet, salty, sour, bitter, umami).

### Chemosensory Perception Test (CPT)

Finally, we remotely administered the CPT. This olfactory test requires participants to smell specific household substances (peanut butter, jam, coffee) and report the perceived intensity of each on a 10-point VAS^20^. Scores were obtained by calculating the mean score reported for the 3 substances. Based on earlier data, we classified participants with a score lower or equal to 7 as exhibiting (c) clinically relevant OD^20^.

#### Statistical Analysis

A custom Python script (Python 3.7.5, Python Software Foundation, https://www.python.org) and SPSS 26.0 (Armonk, NY: IBM Corp) were used to process raw questionnaire data. The Python script was used to merge results from the two questionnaires and SPSS Syntax was used to transform raw data. Processed data were analyzed and visualized with SPSS 26.0, GraphPad Prism 8.3.1 (GraphPad Prism Software, San Diego, CA) and Raincloud plots ^23^.

Student’s T-tests and chi square tests were performed to assess change in (a) chemosensory sensitivity, prevalence of (b) persistent reduction of chemosensory sensitivity and of (c) clinically relevant OD from T2 to T3. To quantify the effects of COVID-19 on *chemosensory modality* (olfactory, gustatory, and trigeminal sensitivity) and *time point* (T0, T1, T2, T3), repeated measures (rm) ANOVA with age as a covariate were computed. To disentangle interactions, separate rmANOVA were carried out for individual chemosensory modalities and at T3 for the same factors. Greenhouse-Geisser corrections were used for sphericity and Tukey’s multiple comparisons tests were performed for post-hoc comparisons.

To determine differentiating characteristics between patients with and without impairment of chemosensory function at T3 after COVID-19 among those with reduction of chemosensory sensitivity during COVID-19, a forward selection logistic regression was performed to ascertain the effects of (1) age, (2) sex, (3) self-rated olfactory function at T0, (4) COVID-19 symptoms at T1 (fever, cough, dyspnea, chest pain, rhinorrhea, changes in food flavour, appetite loss, headache, myalgia, fatigue, diarrhea, abdominal pain, nausea), (5) chronic comorbidities with a prevalence of more than 5% in our sample (hypertension, obesity) and (6) qualitative OD (parosmia, phantosmia, waxing and waning) at T2, on the likelihood of impairment of chemosensory function at T3. Other chronic comorbidities were not included in the model due to presence of these conditions in less than 5% of our population (heart disease, diabetes mellitus, pulmonary disease, neurological disease, cancer). Self-rated olfactory function at T1 was also excluded from the model since it violated the assumption of linearity of the log odds.

Finally, subjective scales have been used widely in questionnaires to quantify the degree of COVID-19-related OD, yet few studies have analyzed the reliability of such measures in the context of the COVID-19 pandemic^8, 24-26^. Measures of chemosensory sensitivity from T0 to T2 were repeated at the 5-month and 11-month questionnaires and compared to determine the test-retest reliability of self-evaluation (complete data set for 276 participants). To assess the intra-rater reliability of self-reported chemosensory ratings, intraclass correlation coefficients (ICC) and their 95% confidence intervals (CI) were calculated based on a mean rating (k=2), absolute agreement, two-way mixed-effects model. The ICC ranges from 0.00 (absence of reliability) to 1.00 (perfect reliability). No standard values exist for acceptable reliability using ICC, but generally, values below 0.50 indicate poor reliability, between 0.50 and 0.75 indicate moderate reliability, between 0.75 and 0.90 indicate good reliability and above 0.90 is excellent reliability ^27^.

For all statistical tests, alpha type error threshold was set at 0.05. All results are expressed as mean and standard deviation (SD) unless otherwise specified.

#### Data availability

Anonymized data not published within this article will be made available by request from any qualified investigator.

## Results

### Study population

A total of 366 healthcare workers were included in this study. The average age was of participants was of 44.8 years old (SD: 11.7). Among them, 310 (84.7%) were women, and the majority (83.1%) were Caucasian. On average, the online questionnaire was completed 10.6 months (SD: 0.7, range: 8.9 - 13.0) after the onset of COVID-19 symptoms. During the acute SARS-CoV-2 infection (T1), 307 (83.9%) and 301 (82.2%) reported lower olfactory and gustatory scores respectively compared to T0. Our study population had a higher prevalence of OD and GD than the initial survey from which our participants were recruited (OD: (χ^2^(1, N= 4908) = 52.62, P < .001); GD: (χ^2^(1, N= 4908) = 68.03, P < .001)) (Table 1).

**Table 1.** Self-reported and semi-objectively measured chemosensory alterations. T1: during SARS-CoV-2 infection; T2: approximately 5 months after SARS-CoV-2 infection; T3: approximately 11 months after SARS-CoV-2 infection; Q1: Questionnaire, sent at T2; Q2: questionnaire sent at T3; OD: Olfactory dysfunction; GD: Gustatory dysfunction.; CPT: Chemosensory Perception Test; INSPQ: Institut National de santé publique du Québec.

|  | Prevalence of self-reported OD in INSPQ study n (%) | Prevalence of self-reported GD in INSPQ study n (%) | Reduced olfactory sensitivity n (%) |  | Reduced gustatory sensitivity n (%) |  | Reduced trigeminal sensitivity n (%) |  | OD measured by CPT n (%) | Impaired olfactory function n (%) | Impaired gustatory function n (%) | Impaired trigeminal function n (%) |
| --- | --- | --- | --- | --- | --- | --- | --- | --- | --- | --- | --- | --- |
|  |  |  | Q1 | Q2 | Q1 | Q2 | Q1 | Q2 |  |  |  |  |
| N | 4542 |  | 366 |  |  |  |  |  |  |  |  |  |
| T1 | 2966 (65.3) | 2748 (60.5) | 307 (83.9) | 306 (83.6) | 301 (82.2) | 297 (81.1) | 175 (47.8) | 196 (53.6) |  |  |  |  |
| T2 |  |  | 155 (56.2) * | 242 (66.1) | 123 (44.6) * | 224 (61.2) | 66 (23.9) * | 133 (36.3) | 108 (29.5) |  |  |  |
| T3 |  |  |  | 184 (50.3) |  | 163 (44.5) |  | 86 (23.5) | 91 (24.9) | 118 (32.2) | 111 (30.3) | 52 (14.2) |

\*n=276

### Self-rated chemosensory sensitivity

Average self-evaluated olfactory sensitivity at T3 was 7.6 (2.2), compared to 9.1 (1.1) at T0; a total of 184 participants (50.3%) indicated persistent reduction of olfactory sensitivity. Average self-evaluated gustatory sensitivity was 8.0 (2.0) at T3, compared to 9.2 (1.5) at T0; a total of 163 participants (44.5%) indicated persistent reduction of gustatory sensitivity. Average self-evaluated trigeminal sensitivity was 8.7 (1.7) at T3, compared to 9.0 (1.7) at T0; a total of 86 (23.5%) indicated persistent reduction of trigeminal sensitivity.

Among 276 participants who had provided self-ratings at T2 and T3, the proportion of participants with persistent reduction of olfactory, gustatory, or trigeminal sensitivity did not change between T2 and T3 (olfaction: χ2(1, N=276) = 2.62, P = .11; gustation: χ2(1, N=276) = 0.007, P = .93; trigeminal: (χ2(1, N=276) = 0.01, P= 0.92).). However, olfactory (t (275) = -3.91, P < .001, 11 months > 5 months), but not gustatory (t (275) = -.673, P = .501) or trigeminal (t (275) = -.798, P = .425) scores increased from 5 months to 11 months (Figure 2).

**Figure 2.**
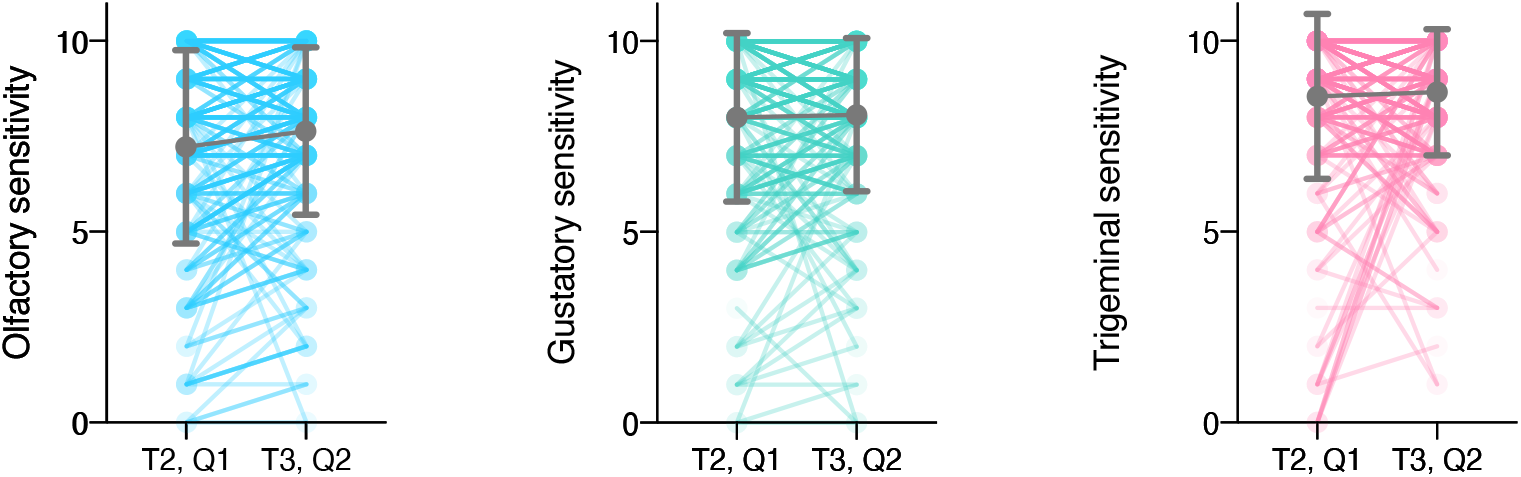
Tendency plots for self-reported chemosensory ratings at 5 months and 11 months following COVID-19 infection. Comparing self-reported chemosensory scores collected at the 5- and 11-month questionnaires. T2, Q1: 5-month rating at the 5-month questionnaire; T3, Q2: 11-month rating at the 11-month questionnaire.

In fact, *chemosensory modality* (F (2, 726) = 24.571, P < .001, η^2^ = .063; olfactory scores < gustatory scores < trigeminal scores; all *P* < .001) and *time point* (F(3, 1089) = 48.88, P < .001, η^2^ = .119; T1 scores < T2 scores < T3 scores < T0 scores, all P < .001) had an effect on self-ratings (Figure 4). More specifically, the effect of *time point* was the strongest for olfactory ratings (F (3, 1089) = 61.677, P <.001 η^2^ = .145; T1 < T2 < T3 < T0, all P < .001), followed by gustatory ratings (F (3, 1089) = 48.654, P <.001 η^2^ = .118; T1 < T2 < T3 < T0, all P < .001), indicating that these two modalities evolve the most in time. For the trigeminal function, *time point* also influenced self-ratings, but the average self-rating at T3 was comparable to that of T0 (F (3, 1089) = 3.506, P = .028 η^2^ = .010; T1< T2< T3 (all previous P < .001) = T0 (T3 vs T0: P =.060)), indicating a return to baseline trigeminal function.

**Figure 3.**
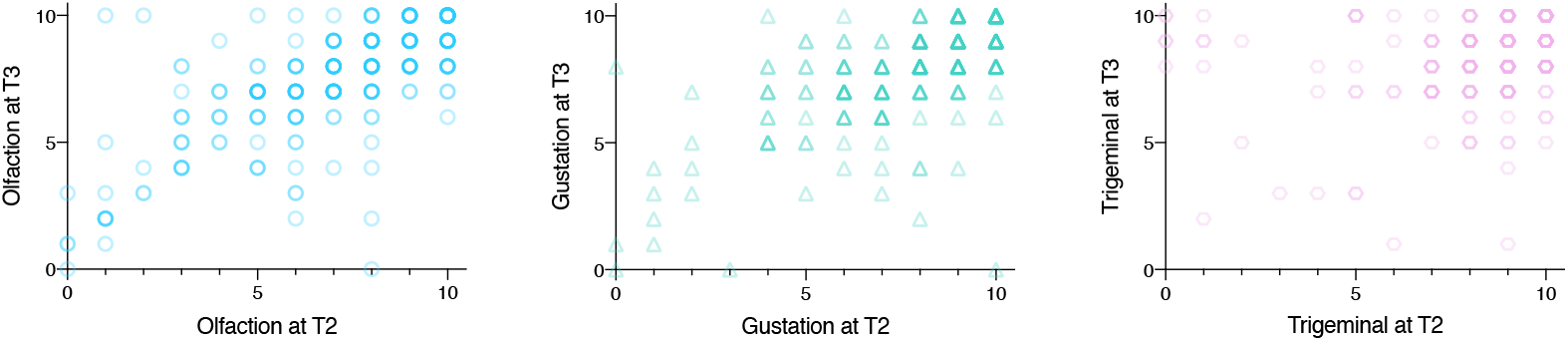
Correlation plots for subjective ratings taken at 5 months and 11 months (n= 276). Correlations between alterations in olfaction (blue circles; ρ = .721, P < .001), gustation (green triangles; ρ = .681 P < .001), and trigeminal function (red hexagons; ρ = .441, P < .001) at 5 and 11 months after infection. Darker colors indicate higher occurrence. T2: 5 months post-COVID-19. T3: 11 months post-COVID-19

**Figure 4.**
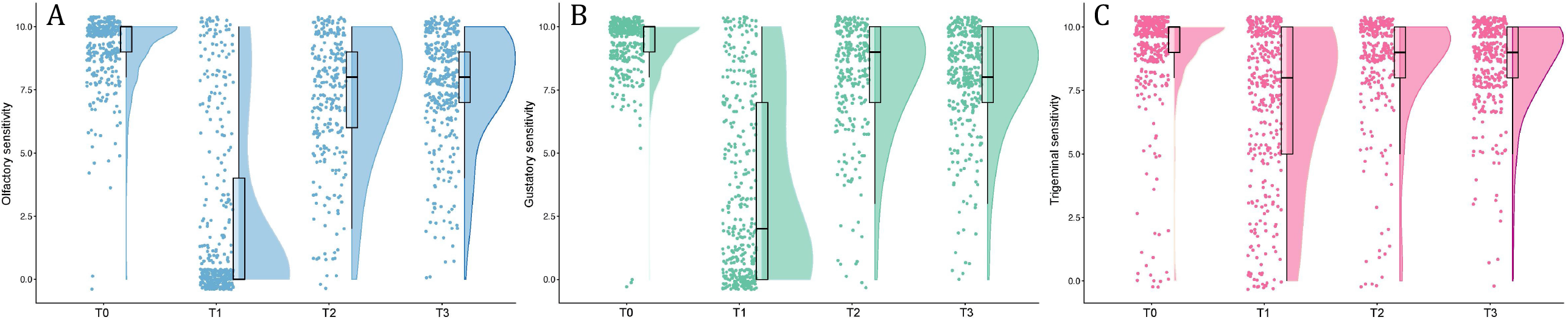
Density distributions of self-reported chemosensory ratings (n= 366). Raincloud plot representing self-reported scores for olfaction (A), gustation (B), and trigeminal (C) function before (T0), during (T1) and 5 (T2) and 11 (T3) months after COVID-19. Ratings from individual participants are displayed as dots. Boxplots show the first to third quartiles, horizontal line denotes the median, and whiskers denote 1.5 times interquartile range. Compared to baseline, self-reported scores of olfaction, gustation and trigeminal function were significantly lower during COVID-19 and have not fully returned to baseline values 11 months after COVID-19.

### Subjective impairment of chemosensory function

When asked to compare T3 with T0, 118 (32.2%; 50 (13.7 %): much worse; 68 (18.6%): a bit worse) participants reported persistent impairment of olfactory function, 111 (30.3%) (29 (7.9%): much worse; 82 (22.4%): a bit worse) persistent impairment of gustatory function and 52 (14.2%) (9 (2.5 %): much worse; 43 (11.7%): a bit worse) reported persistent impairment of trigeminal function (Figure 5).

Among 118 participants with persistent impairment of olfactory function, a total of 68 (56.7%), 33 (28.0%) and 27 (22.9%) reported parosmia, phantosmia and waxing and waning, respectively at T3. One participant reported persistent nasal congestion, while 2 participants could not describe the qualitative impairment. At T2, 20 (16.9%), 27 (22.9%) and 21 (17.8%) had reported parosmia, phantosmia and waxing and waning, respectively.

Among the 111 participants who reported worsened gustatory function at T3, bitter taste was the most affected quality (46.8%), followed by sweet (40.5%), salty (38.7%), umami (33.3%), and sour (32.4%). Among them, 9.9% report a combined gustatory dysfunction involving all 5 taste qualities.

### Clinically relevant olfactory dysfunction

A total of 91 (24.9%) participants exhibited CPT scores equal or less than 7 indicating clinically relevant OD at T3. Although this number was nominally down from 108 (29.5%) at T2, there was no difference in the proportions between both time points (χ^2^ (1, N = 366) = 1.93, *P* = .16). Nevertheless, the average CPT score significantly increased by 0.45 points from T2 (6.9 points) to T3 (7.3 points).

### Association between measures

#### Prediction of persistent impairment of olfactory function

The logistic regression model was statistically (χ^2^(3, N= 307) = 30.77, P < .001) to predict persistent impairment of olfactory function; sensitivity and specificity of the model were respectively of 0.92 and 0.21. Three variables were significantly associated with a higher likelihood of persistent impairment of olfactory function 11 months after COVID-19 (Table 2), namely (1) chest tightness at T1 (during infection), (2) dysgeusia at T1 (during infection), and (3) presence of phantosmia at T2 (5 months after infection).

**Table 2.** Logistic regression model predicting persisting olfactory dysfunction at 11 months following COVID-19) (n= 307).

|  | Variable | b | P | 95% C.I. for Odds Ratio |  |  |
| --- | --- | --- | --- | --- | --- | --- |
|  |  |  |  | Lower | Odds Ratio | Upper |
| Variables included | Chest pain during COVID-19 | 0.563 | <.001 | 1.05 | 1.76 | 2.94 |
|  | Dysgeusia during COVID-19 | 0.856 | .007 | 1.26 | 2.35 | 4.39 |
|  | Phantosmia at 5 months | 0.754 | .030 | 1.08 | 2.13 | 4.20 |
| Variables excluded | Sex | - | 0.789 | - | - | - |
|  | Olfactory sensitivity rating before COVID-19 | - | 0.553 | - | - | - |
|  | Participant age | - | 0.755 | - | - | - |
|  | Fever during COVID-19 | - | 0.299 | - | - | - |
|  | Dry cough during COVID-19 | - | 0.214 | - | - | - |
|  | Productive cough during COVID-19 | - | 0.933 | - | - | - |
|  | Dyspnea during COVID-19 | - | 0.405 | - | - | - |
|  | Rhinorrhea during COVID-19 | - | 0.520 | - | - | - |
|  | Odynophagia during COVID-19 | - | 0.338 | - | - | - |
|  | Appetite loss during COVID-19 | - | 0.923 | - | - | - |
|  | Headache during COVID-19 | - | 0.447 | - | - | - |
|  | Myalgia during COVID-19 | - | 0.111 | - | - | - |
|  | Fatigue during COVID-19 | - | 0.928 | - | - | - |
|  | Diarrhea during COVID-19 | - | 0.326 | - | - | - |
|  | Abdominal pain during COVID-19 | - | 0.211 | - | - | - |
|  | Nausea during COVID-19 | - | 0.448 | - | - | - |
|  | Parosmia at 5 months | - | 0.707 | - | - | - |
|  | Hypertension | - | 0.510 | - | - | - |
|  | Obesity | - | 0.447 | - | - | - |

### Test-retest reliability of subjective chemosensory ratings

Self-ratings from the first and second questionnaires correlated significantly at each of three repeated time points (T0, T1, and T2), and olfaction had the highest correlation coefficients. Notably, for olfaction, the average-measures of the ICC were 0.635 (95 % CI: 0.552 - 0.703), 0.927 (0.908 - 0.941) and 0.818 (95% CI: 0.760 - 0.860), respectively, for T0, T1 and T2 indicating high test-retest reliability. Gustation (T0: 0.332 (0.180 – 0.456); T1: 0.809 (0.865 - 0.910); T2: 0.661 (0.494 - 0.764)) and trigeminal function (T0: 0.388 (0.248 - 0.502); T1: 0.607 (0.515 - 0.681); T2: 0.320 (0.143 - 0.461)) had somewhat lower test-retest reliability (Table 3).

**Table 3.** Intraclass correlation coefficients and Spearman’s correlation coefficients for olfactory, gustatory, and trigeminal subjective ratings at 5 and 11 months.

|  | Olfaction |  | Gustation |  | Trigeminal |  |
| --- | --- | --- | --- | --- | --- | --- |
|  | ICC* (95% CI) | Spearman's | ICC* (95% CI) | Spearman's | ICC* (95% CI) | Spearman's |
| T0<br>Before<br>COVID<br>(n=366) | 0.635 (0.552 -<br>0.703) | 0.577** | 0.332 (0.180<br>- 0.456) | 0.539** | 0.388 (0.248<br>- 0.502) | 0.379** |
| T1<br>During<br>COVID<br>(n=366) | 0.927 (0.908 -<br>0.941) | 0.816** | 0.809 (0.865<br>- 0.910) | 0.796** | 0.607 (0.515<br>- 0.681) | 0.446** |
| T2<br>At 5<br>months<br>(n= 276) | 0.818 (0.760 -<br>0.860) | 0.701** | 0.661 (0.494<br>- 0.764) | 0.579** | 0.320 (0.143<br>- 0.461) | 0.346** |

\*Two-way mixed effects, absolute agreement, single rater
\*\* P < .001

## Discussion

This study was carried out 11 months after RT-PCR-confirmed SARS-CoV-2 infection and revealed three major results. First, we found that a considerable proportion of participants still exhibited chemosensory loss of different degrees. Specifically, (a) we observed persistent reduction of olfactory (gustatory, trigeminal) sensitivity in 50% (45%, 24%) of participants; i.e., ratings had not yet returned to levels before the infection; (b) roughly a third of participants exhibited persistent olfactory and gustatory impairment; this was one out of seven for the trigeminal system; (c) roughly one quarter of participants had test scores indicative of clinically relevant OD. Second, we observed that presence of (a) chest tightness and (b) subjective dysgeusia during COVID-19 as well as (c) presence of phantosmia at 5 months were predictors for persistent olfactory impairment at 11 months. Third, we observed that the measures we used exhibited good test-retest reliability, especially for olfactory measures.

Our sample roughly exhibited 20% higher rates of reduced olfactory and sensitivity during the acute phase of COVID-19 than the initial survey respondents. Therefore, OD and GD prevalence reported from this study are probably overestimated, but it would be difficult to determine exactly how the recovery rates might differ from participating and non-participating individuals. 32% of participants exhibited persistent OD and 25% exhibited a CPT score indicating clinical OD. The prevalence of parosmia in individuals with post-COVID-19 OD has been reported to be in the range of 7% to 93% in varying degrees of severity^8, 13^. In this cohort, prevalence of parosmia was at 17% at 5 months after the infection and rose to over 50% at 11 months^20^. This is in line with a recent study that reported increasing incidence of parosmia in individuals recovering from long-haul COVID-19^17^. This finding is particularly important, as parosmia has been associated with a better olfactory outcome following olfactory training, but if unresolved, it may be a much greater source of distress than isolated hyposmia^28, 29^. Further, an important proportion of COVID-19 patients also reported phantosmia and waxing and waning of olfactory function, although these chemosensory dysfunctions have not been associated with a better outcome^30^. Among all participants, olfactory function measured with a semi-objective test nominally increased between 5 months and 11 months; yet no significant difference was found in OD prevalence at 5 and 11 months using this tool. This could be interpreted as an increase in qualitative disorders combined with improved olfactory sensitivity, or lower spontaneous recovery rates for those with more severe deficits while others with milder and clinically irrelevant forms continue to recuperate.

We found that 30% of the participants in the cohort exhibited persistent GD, with bitter as the most affected taste quality. One study on taste thresholds in COVID-19 patients found that the threshold was increased for sweet, sour and bitter, but they were decreased for salty in the acute phase^31^. TD at 11 months was reported by less than 15% of participants. According to a recent study, impaired trigeminal function could play a role in local inflammatory response, which may in turn influence recovery ^22^.

We explored chemosensory alterations in three different ways: A relatively high number of individuals reported at least slight alterations in chemosensitivity compared to baseline when using VAS ratings. When directly asked if a given chemical senses was a bit worse or much worse compared to baseline, a smaller number of individuals self-reported dysfunction. Therefore, even though a large proportion of individuals with post-COVID-19 chemosensory alterations might feel like their senses are not back to normal, only some of these have more severe dysfunctions. In the clinical setting, it is imperative that clinicians use the same scale or measure if they wish to follow-up subjective olfactory complaints as the answer may vary depending on the formulation of the question.

Few clinical measures proved to be good indicators of olfactory prognostic following COVID-19-induced OD, in line with other reports^32, 33^. In our study, presence of (a) chest tightness and (b) taste disorders during acute COVID-19 as well as (c) presence of phantosmia at 5 months were significantly associated with a higher chance of persisting OD at 11 months. The links between these variables remain speculative. Chest tightness may indicate a more severe form of COVID-19, although OD was not associated with a more severe course of COVID-19^34^. Participants may mistake lack of flavor perception due to reduced retronasal olfaction caused by OD as taste problems in the acute phase of COVID-19^35-37^. This finding could either be due to more severe olfactory loss being related to longer duration of the symptom, or that GD does influence olfactory recovery by unknown mechanisms. Finally, although the pathomechanism of phantosmia is still unknown, one could expect that a more severe inflammatory reaction in the olfactory cleft may cause phantosmia and could take more time to recover^38^. One study found patients with lower levels of salivary and nasal immunoglobulins G at 60 days post-infections had better outcomes^32^. Accordingly, the presence of phantosmia increased the likelihood of OD persistence. While this suggests a role of immune local response in the evolution of persistent chemosensory disorders, further research must be conducted to accurately identify patients with a better chance of olfactory recovery. More studies will be required to validate the link between these variables, especially between qualitative OD and olfactory recovery. Meanwhile, clinicians can only continue to encourage their patients with persisting OD to try olfactory training and/or intranasal corticosteroid sprays following resolution of the infection^39-41^.

Olfactory subjective ratings had a very good test-retest reliability in our study population. Psychophysical evaluations do remain the ideal method to measure all chemosensory disorders but the use of a 10-point VAS may have some value in monitoring OD, especially in large populations or in a social distancing setting. Previous studies have found self-ratings of olfactory function to have a poor reliability and low correlation with objective testing^42^. However, our findings suggest self-ratings could be a good alternative for clinicians with limited time or a difficult access to objective testing. These findings can be explained in the context of follow-up, especially following COVID-19, where patients are more likely to notice changes in their sense of smell through increased self-awareness of symptoms. A previous study also reported a good correlation between VAS-reported OD and the BSIT, a validated psychophysical olfactory test^26^. In our study, gustatory ratings had a moderate test-retest reliability and trigeminal had a poor test-retest reliability. This difference between chemosensory modalities was probably influenced by how well individuals understand the relation between the different sensations and each chemosensory modality. Indeed, the trigeminal system is a much less known system, and gustation is often mixed with retronasal olfaction^43, 44^. In consequence, despite providing specific definitions and examples for all three chemosensory modalities prior to rating, awareness and accuracy were lower for gustation and trigeminal sensations, possibly demonstrating the need for patient’s education when presenting with chemosensory disorders.

### Limitations

A major limitation of this study was the absence of a commonly used psychophysical test to assess the extent of chemosensory dysfunction. Therefore, our results obtained with the CPT were more difficult to compare directly with other studies using psychophysical testing. Unfortunately, in clinical practice, psychophysical evaluations are seldom used for the diagnosis and follow-up of chemosensory impairments, which are mostly treated based on subjective assessments. While we encourage the use of validated psychophysical tests in primary care, neurology and ENT clinics, findings in this study confirmed a reliability of questioning olfactory functions subjectively especially in the context of follow-up of COVID-19 patients. An additional limitation of this study was the high prevalence of chemosensory disorders during the acute phase of COVID-19. Indeed, we observed higher participation rates among individuals with chemosensory disorders than those who have either recovered or simply never noticed any OD, GD or TD. Our study sample had a 20% higher rate of OD and GD, thus probably over estimating prevalence of chemosensory disorders at 11 months.

## Conclusion

In this study, there was limited improvement from 5 to 11 months after COVID-19 infection with a third of patients reporting persistent chemosensory dysfunctions. Prevalence of parosmia and phantosmia increased significantly from 5 to 11 months post-infection, possibly indicating changes in the olfactory epithelium. More studies on the physiopathology underlying post-COVID-19 OD are necessary to develop better treatments and interventions for the patients with persisting chemosensory dysfunctions.

## Supporting information

Appendix 1 : Online Questionnaire

## Data Availability

All data produced in the present study are available upon reasonable request to the authors.

## Acknowledgements

We thank Josiane Rivard for preparing the online questionnaire, study participants. This work was supported financially by Fonds de recherche du Québec – Santé (chercheur boursier junior 2 #283144 to JF). All authors have full access to all the data in the study and take responsibility for the integrity of the data and the accuracy of the data analysis. FGL is the recipient of a tier-2 Canada research Chair. No author declares any conflict of interest.

## Notes

### Competing Interest Statement

The authors have declared no competing interest.

### Funding Statement

This work was supported financially by Fonds de recherche du Quebec, Sante (chercheur boursier junior 2 #283144 to JF).

### Author Declarations

Ethics committee of the CHU de Quebec Universite Laval gave ethical approval for this work (MP 20 2021 5228).

