## Appendix 1 : Online Questionnaire for "Persisting Chemosensory Impairments in 366 Healthcare Workers Following COVID-19: An 11-Month Follow-up"

Supplementary file 1:

**Subjective Chemosensory Dysfunction Questionnaire (SCD-Q, T3)**

| | |
|---|---|
| **D1** | Hello,<br><br>Thank you for agreeing to continue this study with us.<br><br>We offer you a 35 minutes ONLINE QUESTIONNAIRE including a behavioral olfactory test. This AT-HOME SCENT TEST involves smelling and tasting a variety of common substances found in your kitchen to establish your odor detection threshold. |
| **D2** | **Please prepare peanut butter, jam or jelly, vinegar, coffee, salt and sugar if you have any.**<br>**You will need these ingredients for the last section of the survey.** |
| Q1 | **Please select the category that best describes you?**<br><br>Caucasian (White)<br>Indigenous (First Nations, Inuit, Metis)<br>Asian<br>Black<br>Latin-American<br>Arab<br>Other **[text box]**<br>Prefer not to answer |
| Q2 | Have you been **reinfected** with COVID-19?<br><br>-Yes<br><br>-No |
| Q3 | Were you diagnosed with any **other** respiratory illnesses (**not COVID-19**) in the **last two weeks**? (Select all that apply)<br><br>- Strep throat (Streptococcal bacteria)<br>- Another bacterial illness<br>- Flu (influenza)<br>- Another viral illness<br>- Other<br>- **None** [**Exclusive]** |
| Q4 | Have you had any of the following symptoms with your **recent respiratory illness or diagnosis (NOT COVID)** (Select all that |

| | |
|---|---|
| | apply)<br><br>- Fever<br>- Dry cough<br>- Cough with mucus<br>- Difficulty breathing / shortness of breath<br>- Chest tightness<br>- Runny nose<br>- Sore throat<br>- Changes in food flavor<br>- Changes in smell<br>- Loss of appetite<br>- Headache<br>- Muscle aches<br>- Fatigue<br>- Diarrhea<br>- Abdominal pain<br>- Nausea<br>- No symptoms **[Exclusive]** |
| Q5 | What treatment(s) or medication(s) have you received for your recent respiratory illness or diagnosis (NOT COVID)?<br><br>- **[text box]**<br>- None |
| **D3** | **The next question relates to your pre-existing medical conditions.** |
| Q6 | Have you been treated or diagnosed for any of the following? (Select all that apply)<br>- High blood pressure<br>- Heart disease (heart attack or stroke)<br>- Diabetes (high blood sugar)<br>- Obesity<br>- Lung disease (asthma/chronic bronchitis)<br>- Head trauma<br>- Neurological disease<br>- Cancer that required chemotherapy or radiation<br>- Cancer that did NOT require chemotherapy or radiation<br>- Chronic sinus disease<br>- Seasonal allergies / hay fever<br>- Other [text box]<br>- None **[Exclusive]**<br>- Prefer not to answer |
| D5 | **The following questions relate to your <u>sense of smell</u> (for example, sniffing flowers or soap, or smelling garbage) but <u>not the flavor of food</u> in your mouth.** |
| Q7 | Rate your ability to **<u>smell</u>** BEFORE the COVID-19 pandemic. |

| | |
|---|---|
| | 0: No sense of smell …<br>10: Excellent sense of smell |
| Q8 | Rate your ability to **smell** DURING your most recent **COVID-19 infection or diagnosis.**<br><br>0: No sense of smell …<br>10: Excellent sense of smell |
| Q9 | Rate your ability to **smell** TODAY.<br><br>0: No sense of smell …<br>10: Excellent sense of smell |
| Q10 | Rate your ability to **smell** TODAY.<br><br>0: No sense of smell …<br>10: Excellent sense of smell |
| Q11 | Rate your ability to **smell** when you answered our first questionnaire.<br><br>0: No sense of smell …<br>10: Excellent sense of smell<br>Don't remember |
| Q12 | Rate your ability to **smell** COMPARED to the time before the COVID-19 pandemic.<br><br>Much worse<br>A bit worse<br>The same **(skip to D7)**<br>A bit better<br>Much better |
| Q13 | Do you currently experience any changes in your sense of smell? (Select all that apply)<br>- I cannot smell at all / Smells smell less strong than they did before<br>- Smells smell different than they did before (the quality of smell has changed)<br>- I can smell things that aren't there (for example I smell burning when nothing is on fire)<br>- Sense of smell fluctuates (comes and goes)<br>- Other or details to answers selected above **[text box]**<br>- No changes noticed **(Exclusive)** |
| **D4** | **The next questions should be answered spontaneously.**<br>**We will ask you to select one of these 4 answers regarding various statements:** |

| | |
|---|---|
| | Completely agree<br>Partially agree<br>Partially disagree<br>Completely Disagree |
| Q14 | The changes in my sense of smell make me feel isolated.<br><br>Completely agree<br>Partially agree<br>Partially disagree<br>Completely disagree |
| Q15 | Because of the changes in my sense of smell, I have problems with taking part in activities of daily life.<br><br>Completely agree<br>Partially agree<br>Partially disagree<br>Completely Disagree |
| Q16 | The changes in my sense of smell make me feel angry.<br><br>Completely agree<br>Partially agree<br>Partially disagree<br>Completely disagree |
| Q17 | Because of the changes in my sense of smell, I don’t enjoy drinks or food as much as I used to.<br><br>Completely agree<br>Partially agree<br>Partially disagree<br>Completely disagree |
| Q18 | Because of the changes in my sense of smell, I eat less than I used to or more than I used to.<br><br>Completely agree<br>Partially agree<br>Partially disagree<br>Completely disagree |
| Q19 | Because of the changes in my sense of smell, I try harder to relax.<br><br>Completely agree<br>Partially agree<br>Partially disagree<br>Completely disagree |
| Q20 | I am worried that I will never get used to the changes in my sense |

| | |
|---|---|
| | of smell.<br><br>Completely agree<br>Partially agree<br>Partially disagree<br>Completely disagree |
| **D5** | **The following questions relate to the obstruction or congestion of your nose (stuffy nose).** |
| Q21 | How **blocked** was your nose BEFORE the COVID-19 pandemic?<br><br>0: Not at all blocked …<br>10: Completely blocked |
| Q22 | How **blocked** was your nose DURING your most recent **COVID-19 infection or diagnosis**?<br><br>0: Not at all blocked …<br>10: Completely blocked |
| Q23 | How **congested or clogged** is your nose TODAY?<br><br>0: Not blocked at all …<br>10: completely blocked |
| Q24 | How **congested or clogged** was your nose when you answered our first survey?<br><br>0: Not blocked at all …<br>10: completely blocked<br>Don't remember |
| Q25 | Rate how **congested or clogged** your nose is COMPARED to the time before your infection.<br><br>Much more blocked<br>A bit more blocked<br>The same<br>A bit less blocked<br>Much less blocked |
| **D6** | **The following questions are related to your sense of taste. For example, sweetness, sourness, saltiness, bitterness experienced in the mouth.** |
| Q26 | Rate your ability to **taste** BEFORE the COVID-19 pandemic.<br><br>0: No sense of taste …<br>10: Excellent sense of taste |
| Q27 | Rate your ability to **taste** DURING your most recent **COVID-19 infection or diagnosis.** |

| | |
|---|---|
| | 0: No sense of taste …<br>10: Excellent sense of taste |
| Q28 | Rate your ability to **taste** TODAY.<br><br>0: No sense of taste<br>10: excellent sense of taste |
| Q29 | Rate your ability to **taste** when you answered our first questionnaire.<br><br>0: No sense of taste …<br>10: Excellent sense of taste<br>Don't remember |
| Q30 | Rate your ability to **taste** COMPARED to the time before your infection.<br><br>Much worse<br>A bit worse<br>The same<br>A bit better<br>Much better |
| Q31 | Do you currently experience changes with your sense of taste? (Select all that apply)<br>- Sweet (Sugar, caramel, candy, etc.)<br>- Salty (Salt, sea water, smoked or cured meat, etc.)<br>- Sour (Yellow lemon, plain yogurt, etc.)<br>- Bitter (white skin on grapefruit, raw endives, coffee, etc.)<br>- Savory/Umami (ripe tomatoes, soy sauce, parmesan, or cheddar cheese, etc.)<br>- Other or details to answers selected above **[text box]**<br>- No change noticed **[Exclusive]** |
| D7 | **The following questions are related to other sensations in your mouth,like burning, cooling, or tingling. For example, chili peppers, mint gum or candy, or carbonation.** |
| Q32 | Rate your **ability to feel these other sensations** (burning, cooling, or tingling) BEFORE **the COVID-19 pandemic.**<br><br>0: Not sensitive at all….<br>10: Very sensitive |
| Q33 | Rate your **ability to feel these other sensations** (burning, cooling, or tingling) DURING your most recent **COVID-19 infection or diagnosis.**<br><br>0: Not sensitive at all….<br>10: Very sensitive |

| | |
|---|---|
| Q34 | Rate your **ability to feel other sensations like burning, cooling, and tingling** AFTER acute symptoms.<br><br>0: No perception at all…<br>10: very sensitive |
| Q35 | Rate your **ability to feel other sensations like burning, cooling, and tingling** TODAY.<br><br>0: No perception at all…<br>10: very sensitive |
| Q36 | Rate your ability to **ability to feel other sensations like burning, cooling, and tingling** when you answered our first questionnaire.<br><br>0: No perception at all…<br>10: very sensitive |
| Q37 | Rate your **ability to feel other sensations like burning, cooling, and tingling** COMPARED to the time before your infection.<br><br>Much worse<br>A bit worse<br>The same<br>A bit better<br>Much better |
| Q38 | Think about a food or beverage you consume regularly – for example, your morning coffee or tea or a piece of fruit you have each day.<br><br>Has the taste, smell, or flavor changed and not recuperated since your **COVID-19 infection** or diagnosis? If so, **please describe how and be sure to indicate which food or beverage you are describing.**<br><br>**[text box]**<br>No change |
| **D8** | **For the next section, please provide yourself these items from your kitchen:**<br>• **Peanut butter**<br>• **Fruit jam or jelly (Room temperature)**<br>• **Vinegar**<br>• **Coffee (beans, ground, instant, other)**<br>• **Salt**<br>• **Sugar**<br>**2 glasses of lukewarm water** |
| Q39 | Do you have peanut butter?<br><br>Yes<br>No/I’m allergic **(Skip to Q43)** |

| | |
|---|---|
| Q40 | Please **sniff** the peanut butter<br>How strong does the peanut butter smell?<br><br>0: Not at all<br>10: Very strong |
| Q41 | Is the peanut butter smell the same as usual?<br><br>Yes<br>No, weaker<br>No, different |
| Q42 | Without knowing the substance you are going to smell, do you believe you would have recognized the smell of peanut butter?<br><br>Yes<br>No |
| Q43 | Do you have fruit jam or jelly?<br><br>Yes<br>No **(Skip to Q47)** |
| Q44 | Please **sniff** the fruit jam or jelly.<br>How strong does the fruit jam or jelly smell?<br><br>0: Not at all<br>10: Very strong |
| Q45 | Is the fruit jam or jelly smell the same as usual?<br><br>Yes<br>No, weaker<br>No, different |
| Q46 | Without knowing the substance you are going to smell, do you believe you would have recognized the smell of fruit jam or jelly?<br><br>Yes<br>No |
| Q47 | Do you have vinegar?<br><br>Yes<br>No **(Skip to Q51)** |
| Q48 | Please **sniff** the **vinegar**<br>How strong does the vinegar smell?<br><br>0: Not at all<br>10: Very strong |
| Q49 | Is the vinegar smell the same as usual? |

| | |
|---|---|
| | Yes<br>No, weaker<br>No, different |
| Q50 | Without knowing the substance you are going to smell, do you believe you would have recognized the smell of vinegar?<br><br>Yes<br>No |
| Q51 | Do you have coffee (beans, ground, instant, other)?<br><br>Yes<br>No (**<u>skip to D9)</u>** |
| Q52 | Please **sniff** the **coffee** (beans, ground, instant, other)<br><br>How strong does the coffee powder smell?<br><br>0: Not at all<br>10: Very strong |
| Q53 | Is the coffee smell the same as usual?<br><br>Yes<br>No, weaker<br>No, different |
| Q54 | Without knowing the substance you are going to smell, do you believe you would have recognized the smell of coffee?<br><br>Yes<br>No |
| **D9** | **Let's proceed to the tasting section of the test.**<br><br>**(Exclude Q55-57 if no PB and exclude Q58-60 if no jam/jelly)** |
| Q55 | Please put a small amount of **peanut butter** in your mouth. How strong does the peanut butter **taste**?<br><br>0: Not at all<br>10: Very strong |
| Q56 | Does the peanut butter taste the same as usual?<br><br>Yes<br>No, weaker<br>No, different |
| Q57 | Without knowing the substance you are going to taste, do you believe you would have recognized the taste of peanut butter?<br><br>Yes |

| | |
|---|---|
| | No |
| Q58 | Please put a small amount of **fruit jam or jelly** in your mouth. How strong does the fruit jam or jelly **taste**?<br><br>0: Not at all<br>10: Very strong |
| Q59 | Does the fruit jam or jelly taste the same as usual?<br><br>Yes<br>No, weaker<br>No, different |
| Q60 | Without knowing the substance you are going to taste, do you believe you would have recognized the taste of fruit jam or jelly?<br><br>Yes<br>No |
| Q61 | Do you have salt?<br><br>Yes<br>No **(Skip to Q65)** |
| Q62 | Please put a teaspoon of **salt** into a glass filled with 1 cup of lukewarm water. Stir. Then please take a spoonful of water into your mouth. How strong does the salty water **taste**?<br><br>0: Not at all<br>10: Very strong |
| Q63 | Is the salty taste the same as usual?<br><br>Yes<br>No, weaker<br>No, different |
| Q64 | Without knowing the substance you are going to taste, do you believe you would have recognized the salty taste?<br><br>Yes<br>No |
| Q65 | Do you have sugar?<br><br>Yes<br>No **(Skip to Q69)** |
| Q66 | Please put 3 teaspoons of **sugar** into the other glass with 1 cup of lukewarm water. Stir. Then please take a spoonful of water into your mouth. How strong does the sweet water **taste**? |

| | |
|---|---|
| | 0: Not at all<br>10: Very strong |
| Q67 | Is the sweet taste the same as usual?<br><br>Yes<br>No, weaker<br>No, different |
| Q68 | Without knowing the substance you are going to taste, do you believe you would have recognized the sweet taste?<br><br>Yes<br>No |
| Q69 | Would you like to mention anything else regarding your medical condition, the evolution of your chemical senses (smell, taste, other sensations) or any other relevant information?<br><br>Yes [text box]<br>No |
| D10 | Thank you for answering this second-and last online questionnaire! Your contribution to our understanding of this new virus is greatly appreciated.<br><br>Cordially,<br><br>Dr Nicolas Dupré<br>Dr Johannes Frasnelli |
